# Direct medical costs of hospitalized patients with idiopathic pulmonary fibrosis in China

**DOI:** 10.1101/19010025

**Authors:** Xiaofen Zheng, Bingbing Xie, Yan Liu, Ming Zhu, Shu Zhang, Chengjun Ban, Jing Geng, Dingyuan Jiang, Yanhong Ren, Huaping Dai, Chen Wang

## Abstract

**Background:** Idiopathic pulmonary fibrosis (IPF) is a chronic, progressive fibrosing interstitial pneumonia of unknown cause. The incidence of IPF is increasing year by year, as well as the mortality rates, which is really a burden both for the family and the society. However few data concerning the economic burden of the patients with IPF is available, especially in China.

**Objective:** This study aimed to examine the direct medical costs of hospitalized patients with IPF and to determine the contributing factors.

**Methods:** This retrospective analysis used the cost-of-illness framework in order to analyze the direct medical costs of patients with IPF. The study used data from the pneumology department of Beijing Chao-Yang Hospital affiliated to Capital Medical University from year 2012 to 2015. The direct medical costs included drug fee, auxiliary examination fee, treatment fee and other fee. Patients’ characteristics, medical treatment, and the direct medical costs were analyzed by descriptive statistics and multivariable regression.

**Results:** There were 219 hospitalized patients meeting the diagnosis of IPF, 91% male. The mean age was 65 years old. For the direct medical costs of hospitalized patients with IPF, the mean(SD) of the total costs per IPF patient per admission was 14882.3 (30975.8)CNY. The largest parts were the examination fee of 6034.5 (15651.2)CNY and the drug fee of 5048.9 (3855.1)CNY. By regression analysis we found that length of stay, emergency treatment, ventilator use and being a Beijing native were significantly (P<0.05) associated with total hospitalization costs, and the length of stay had the biggest impact. Complications or comorbidities contributated to the direct medical costs as follows: respiratory failure with 30898.3CNY (P=0.004), pulmonary arterial hypertension(PAH) with 26898.2CNY (P=0.098), emphysema with 25368.3CNY (P=0.033), and high blood pressure with 24659.4CNY (P=0.026). Using DLCO or DLCO% pred to reflect the severity of IPF, there was no significant correlation between DLCO or DLCO% pred and patients’ direct medical costs. While, the worse the diffusion function, the higher the drug fee.

**Conclusion:** This study showed that IPF has a major impact on the direct medical costs. Thus, appropriate long-term interventions are recommended to lower the economic burden of IPF.

**Strengths and limitations of this study:** It was the first time in China to discuss the economic burden of diseases and its influencing factors in patients with IPF.

The results of this study might be of reference for the establishment of IPF disease-related medical policies in future.

The retrospective cross-sectional design does not allow for establishing any causal relationships.

It was a a single-center study, resulting a slightly smaller sample size. A large sample of multicenter studies is needed to confirm this.

## Introduction

Idiopathic pulmonary fibrosis (IPF) is defined as a specific form of chronic fibrosing interstitial pneumonia limited to the lung, which is characterized by the pathologic pattern of usual interstitial pneumonia (UIP) and affects usually the elderly. Its etiology is unclear, but genetic factors, smoking and occupational environment exposure may be risk factors.[1-3]

The estimates of the incidence of IPF show a wide variation in different countries, ranging from 3 to 9 per 100000 per year in North America and Europe, while in South America and Asia the incidence was reported to be lower, ranging from 1.2 to 4.16 per 100000 per year.[4] A recent study confirmed that the higher the age of the patient the higher the incidence of IPF.[5] The studies also showed that the mortality was increasing, ranging from 4.68 to 13.36 per 100000. In England and Wales, the deaths from IPF have tripled in the past 20 years.[4]

In China, there are no national population-based data on incidence, prevalence and mortality of IPF. However, data from a large ILD center showed that IPF is the most common subtype of ILD, with increasing numbers of hospitalized patients. With more patients living with IPF, it is important to understand the economic burden associated with this disease.[6]

## Methods

### Design and date sources

This research was designed as a retrospective cross-sectional analysis. Claims data were obtained from the IPF cohort and the database of discharged patients of Beijing Chao-Yang Hospital from the years 2012 to 2015 (219 cases). Data were retrieved from the hospital case statistics management system, including patient characteristics, co-morbid conditions, health-care use and cost. IPF was diagnosed according to the ATS/ERS/JRS/ALAT Statement.[3]Co-morbid conditions included complications and comorbidities, which were defined as any listing of the specific co-morbid diagnosis code until and during the hospitalization. These included respiratory infections, respiratory failure, pulmonary arterial hypertension (PAH), lung cancer, high blood pressure (HBP), coronary heart disease, emphysema, diabetes, gastroesophageal reflux disease (GERD), heart failure, asthma, bronchiectasis. In our study, since all patients lacked right heart catheter monitoring, pulmonary hypertension was defined as an estimated sPAP ≥37 mmHg by Doppler echocardiography based on the 2009 European Society of Cardiology (ESC)/ERS PH Guideline and was divided into three grades:[7] (1) PH unlikely: TRV ≤2.8 m/s, sPAP ≤36 mmHg. (2) PH possible: TRV 2.9– 3.4 m/s, sPAP 37–50 mmHg. (3) PH likely: TRV >3.4 m/s, sPAP >50 mmHg.

The study was reviewed and approved by the Human Ethics Review Committee of the Beijing Chao-Yang Hospital. Written informed consent was obtained from all the patients.

This study used the direct medical costs of hospitalized patient with IPF, which reflect the direct medical economic burden of these patients, covering six categories with a total of 19 cost items (Table 1). The cost estimates were reported in Chinese Yuan (CNY) (1 US dollar ≈ 6.2 CNY, 2015).

**Table 1.**
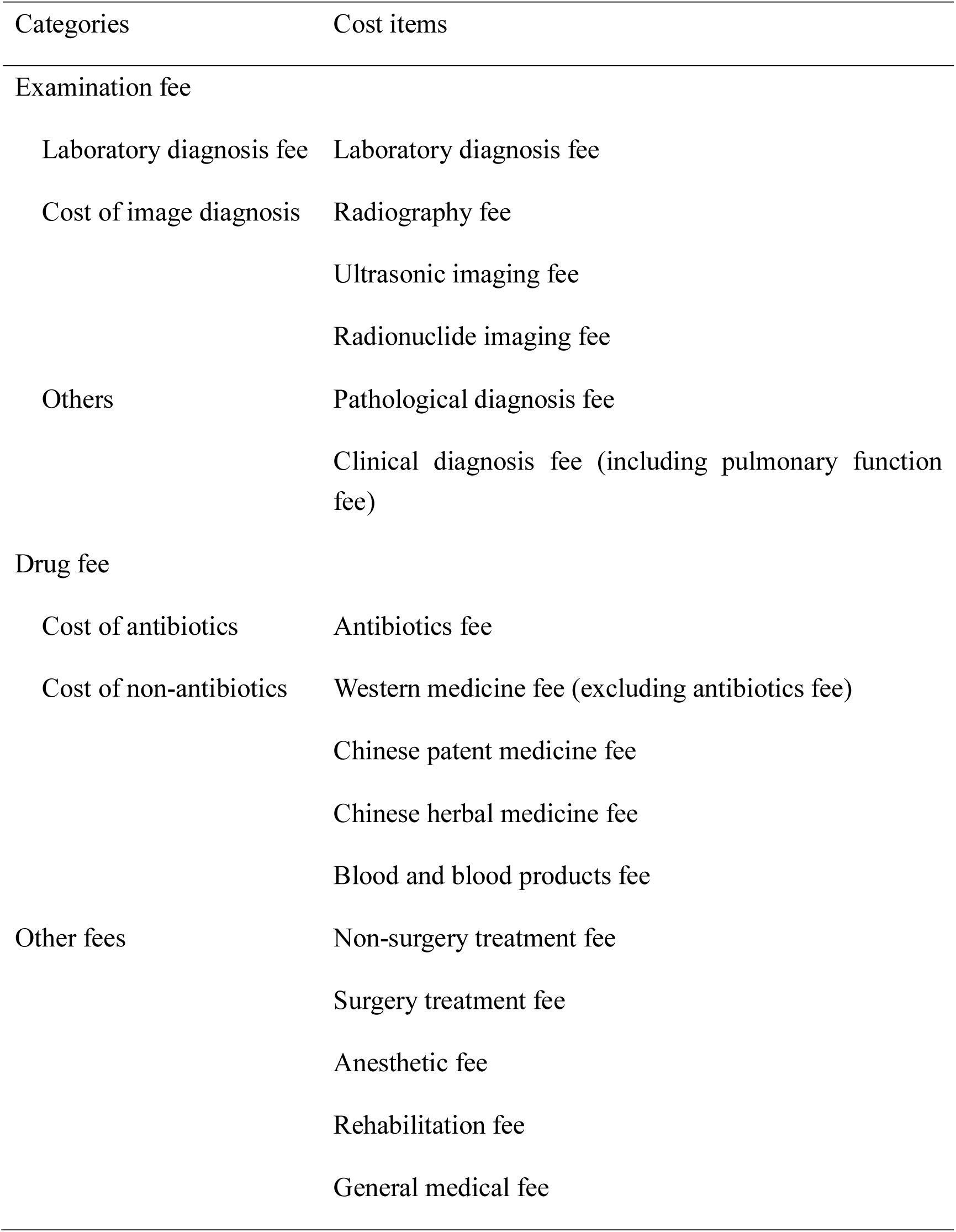
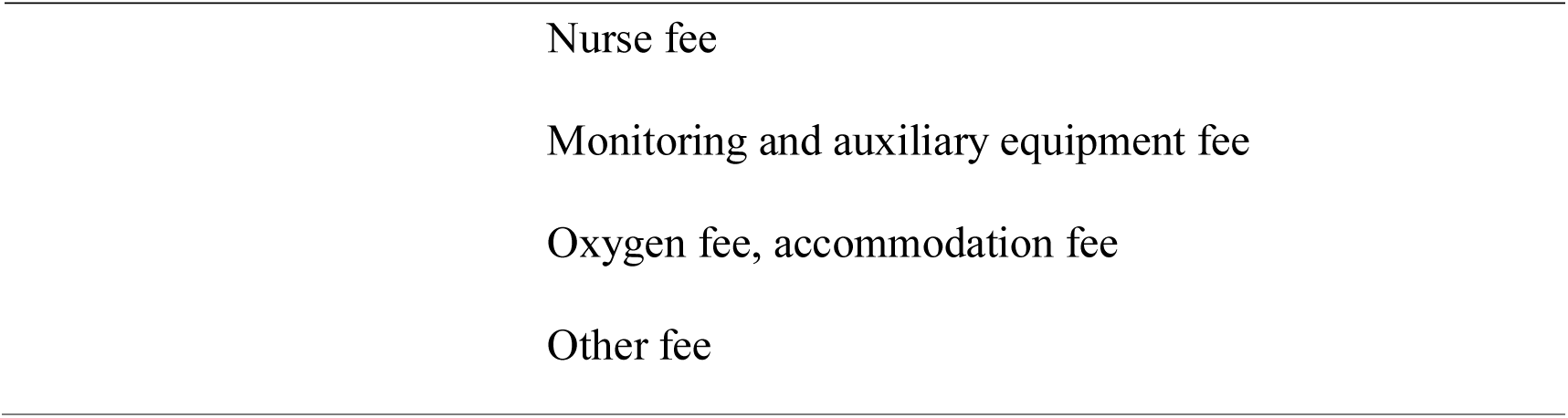
The cost items of the direct medical costs of hospitalized patient with IPF

| Categories | Cost items |
| --- | --- |
| Examination fee |  |
| Laboratory diagnosis fee | Laboratory diagnosis fee |
| Cost of image diagnosis | Radiography fee |
|  | Ultrasonic imaging fee |
|  | Radionuclide imaging fee |
| Others | Pathological diagnosis fee |
|  | Clinical diagnosis fee (including pulmonary function fee) |
| Drug fee |  |
| Cost of antibiotics | Antibiotics fee |
| Cost of non-antibiotics | Western medicine fee (excluding antibiotics fee) |
|  | Chinese patent medicine fee |
|  | Chinese herbal medicine fee |
|  | Blood and blood products fee |
| Other fees | Non-surgery treatment fee |
|  | Surgery treatment fee |
|  | Anesthetic fee |
|  | Rehabilitation fee |
|  | General medical fee |

|  |
| --- |
| Nurse fee |
| Monitoring and auxiliary equipment fee |
| Oxygen fee, accommodation fee |
| Other fee |

### Statistical analysis

Descriptive analyses were conducted to assess HRU (Health Care Resource Utilization) of patients with IPF. Means and standard deviations (SD) were reported for continuous variables, and proportions were reported for categorical variables. The rank-sum test was used to detect the differences for continuous variables. A multiple linear regression (MLR) was utilized to estimate the impact on costs, with the use of step wise regression. Nine independent variables were entered into the regression model, including gender, age, native place reimbursement, length of stay, hospital outcome, ventilator use, and emergency treatment(as shown in Table 2). A generalized linear model (GLM) was used to evaluate which co-morbid conditions drive total costs after accounting for patient characteristics in the total population. P-value < 0.05 was considered statistically significant.

**Table 2.**
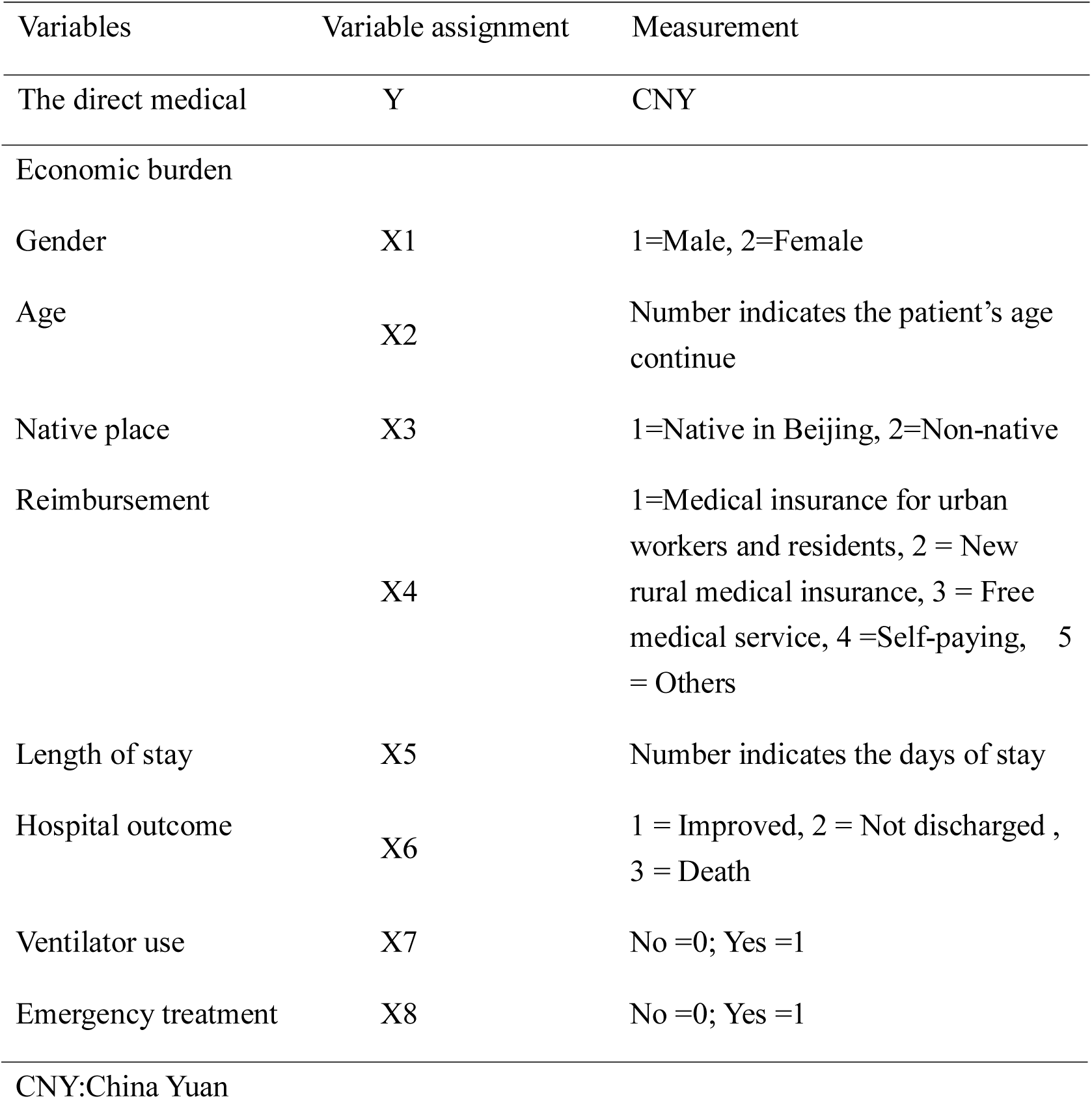
Variables and their measurement in the MLR

| Variables | Variable assignment | Measurement |
| --- | --- | --- |
| The direct medical | Y | CNY |
| Economic burden |  |  |
| Gender | X1 | 1=Male, 2=Female |
| Age | X2 | Number indicates the patient's age<br>continue |
| Native place | X3 | 1=Native in Beijing, 2=Non-native |
| Reimbursement | X4 | 1=Medical insurance for urban<br>workers and residents, 2 = New<br>rural medical insurance, 3 = Free<br>medical service, 4 =Self-paying, 5<br>= Others |
| Length of stay | X5 | Number indicates the days of stay |
| Hospital outcome | X6 | 1 = Improved, 2 = Not discharged ,<br>3 = Death |
| Ventilator use | X7 | No =0; Yes =1 |
| Emergency treatment | X8 | No =0; Yes =1 |

CNY:China Yuan

## Results

### Patient Population

A total of 219 hospitalized patients met the diagnosis of IPF in Beijing Chao-Yang hospital from 2012 to 2015. The patients with IPF were on average 65 years old, 91% male. In terms of age, the youngest patient was 40 years old, while the oldest was 88 years old. Patients aged 61-70 years accounted for 46.6% of the patients. The population was geographically diverse, 48.9% of the patients were from Beijing(Table 3). IPF patients have a variety of co-morbid conditions. The prevalence of selected co-morbid conditions of IPF patients was showed in table 4.

**Table 3.** Baseline demographics and selected healthcare utilization of IPF

| Characteristic | IPF |  |
| --- | --- | --- |
| Subjects, n | 219 | 100% |
| Male | 200 | 91.3% |
| Age, years <sup>a</sup> | 65 | (8) |
| ≤50 | 8 | 3.7% |
| 51-60 | 51 | 23.3% |
| 61-70 | 102 | 46.6% |
| 71-80 | 43 | 19.6% |
| ≥80 | 15 | 6.8% |
| Geographic region |  |  |
| Beijing | 107 | 48.9% |
| HRU |  |  |
| hospitalized days <sup>a</sup> | 10.14 | ( 8 ) |
| chest X-ray | 34 | 16% |
| CT scan | 131 | 60% |
| pulmonary function tests | 131 | 60% |
| bronchoscopy | 94 | 43% |
| oxygen therapy | 184 | 84% |
| mechanical ventilation | 13 | 6% |

IPF, idiopathic pulmonary fibrosis; SD, Standard Deviation; HRU, Health Care Resource Utilization <sup>a</sup> Mean (SD).

**Table 4.** Prevalence of selected co-morbid conditions of IPF patients

| Co-morbid conditions | IPF (n=219) |  |
| --- | --- | --- |
|  | n | % |
| Pulmonary infection | 70 | 32% |
| Respiratory failure | 43 | 19.6% |
| Pulmonary arterial hypertension | 39 | 17.8% |
| Lung cancer | 4 | 1.8% |
| High blood pressure | 56 | 25.6% |
| Coronary artery disease | 41 | 18.7% |
| Emphysema | 35 | 16% |
| Diabetes | 34 | 15.5% |
| GERD* | 16 | 7.3% |
| Heart failure | 9 | 4.1% |
| Asthma | 5 | 2.3% |
| Bronchiectasis | 3 | 1.4% |

IPF, idiopathic pulmonary fibrosis; COPD, chronic obstructive pulmonary disease; GERD, gastroesophageal reflux disease;
\* : 8 cases were diagnosed with 24-hour ambulatory esophageal pH and pressure recording , 8 cases were diagnosed by gastroscopy and upper gastrography.

### Health Care Resource Utilization

The mean(SD) length of hospital stay was 10.1(8) days. Procedures such as chest X-ray (16%), computed tomography (CT) scan (60%), pulmonary function tests (60%), bronchoscopy (43%) and oxygen therapy (84%) were frequently applied. Moreover, 5.9% of cases were treated with invasive or noninvasive ventilation, and 1.4% in the ICU.

### Costs

For hospitalized patients with IPF, the mean (SD) direct medical costs was 14882.3 (30975.8) CNY per capita per admission. The results of costs of medical services for IPF are summarized in Table 5. This table shows that the examination fee, with spending of 6034.5 (15651.2) CNY in total, was the largest proportion (41%) of the direct medical costs. Table 5 also shows that the cost of antibiotics was as much as of non-antibiotics. More details are summarized in Table 5.

**Table 5.**
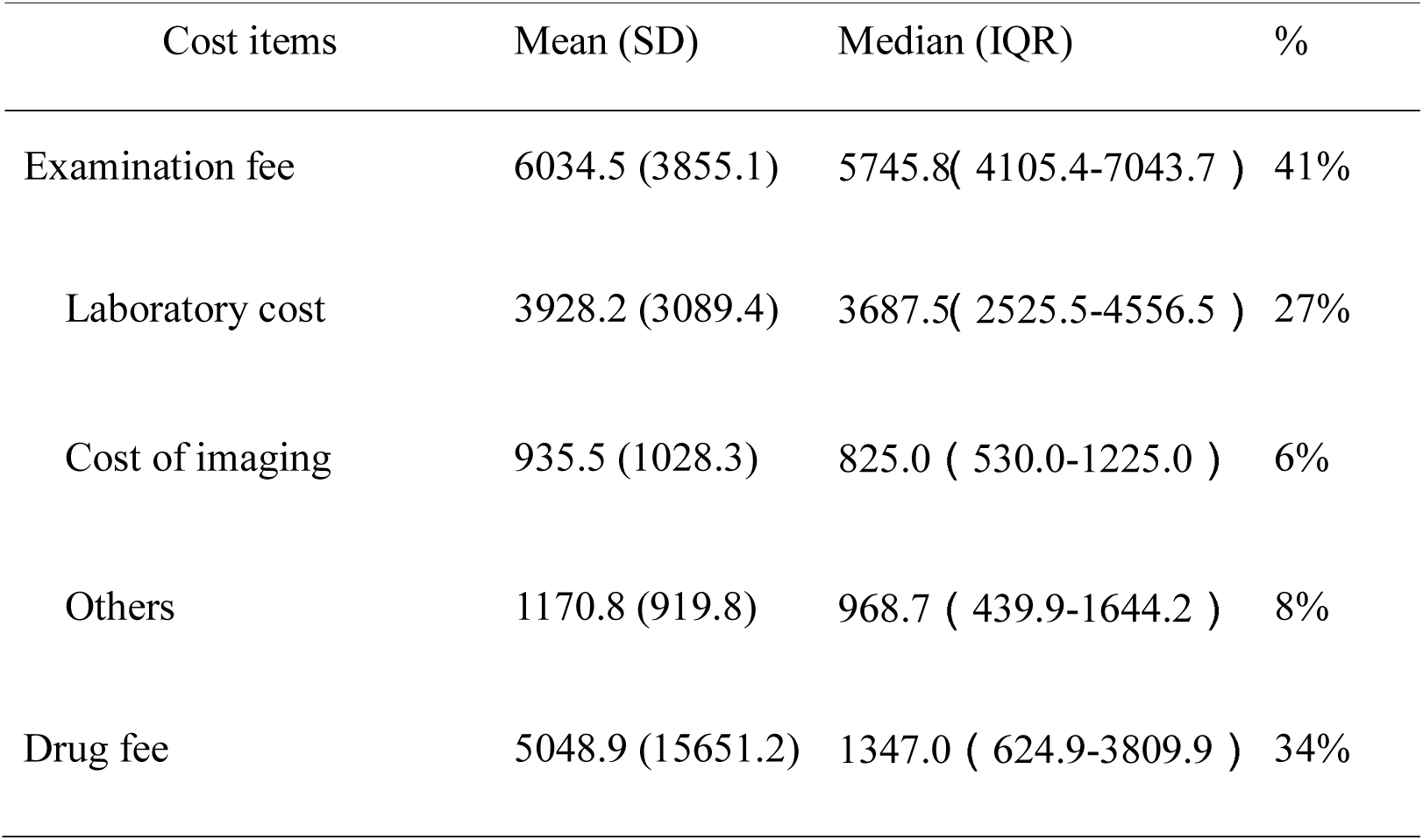
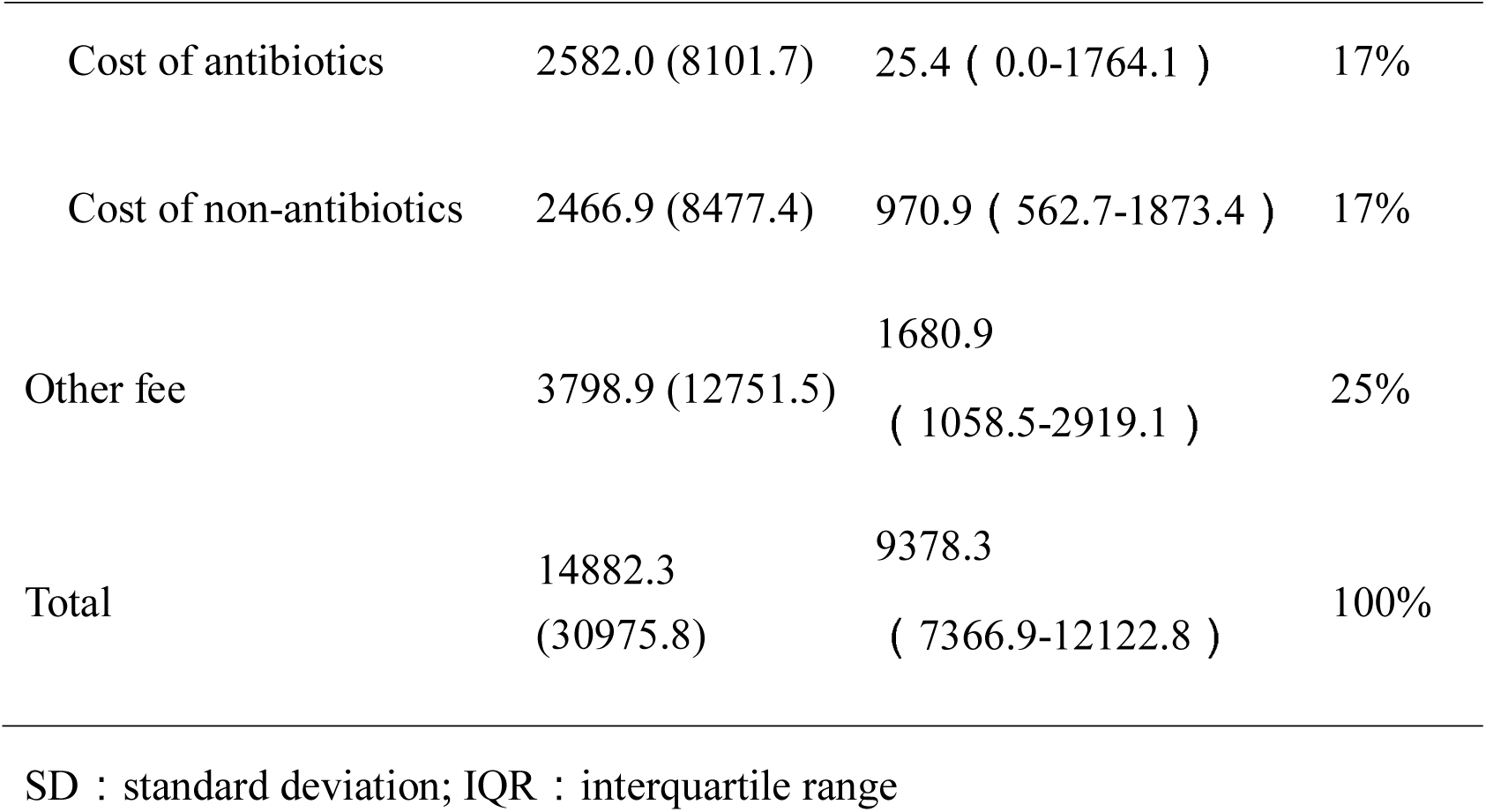
The direct medical costs of hospitalized patients with IPF (CNY, per capita)

| Cost items | Mean (SD) | Median (IQR) | % |
| --- | --- | --- | --- |
| Examination fee | 6034.5 (3855.1) | 5745.8( 4105.4-7043.7 ) | 41% |
| Laboratory cost | 3928.2 (3089.4) | 3687.5( 2525.5-4556.5 ) | 27% |
| Cost of imaging | 935.5 (1028.3) | 825.0 ( 530.0-1225.0 ) | 6% |
| Others | 1170.8 (919.8) | 968.7 ( 439.9-1644.2 ) | 8% |
| Drug fee | 5048.9 (15651.2) | 1347.0 ( 624.9-3809.9 ) | 34% |
| Cost of antibiotics | 2582.0 (8101.7) | 25.4 ( 0.0-1764.1 ) | 17% |
| Cost of non-antibiotics | 2466.9 (8477.4) | 970.9 ( 562.7-1873.4 ) | 17% |
| Other fee | 3798.9 (12751.5) | 1680.9<br>( 1058.5-2919.1 ) | 25% |
| Total | 14882.3<br>(30975.8) | 9378.3<br>( 7366.9-12122.8 ) | 100% |

SD : standard deviation; IQR : interquartile range

The direct medical costs of IPF decreased during the 4 years of this study, from 16219.2CNY in 2012 to 13513.8CNY in 2015 (Fig.1). This was mainly due to a decrease in the drug fee from 5908.0CNY to 3476.6CNY due to the policy of medicine fee decreasing.

**Fig.1.**
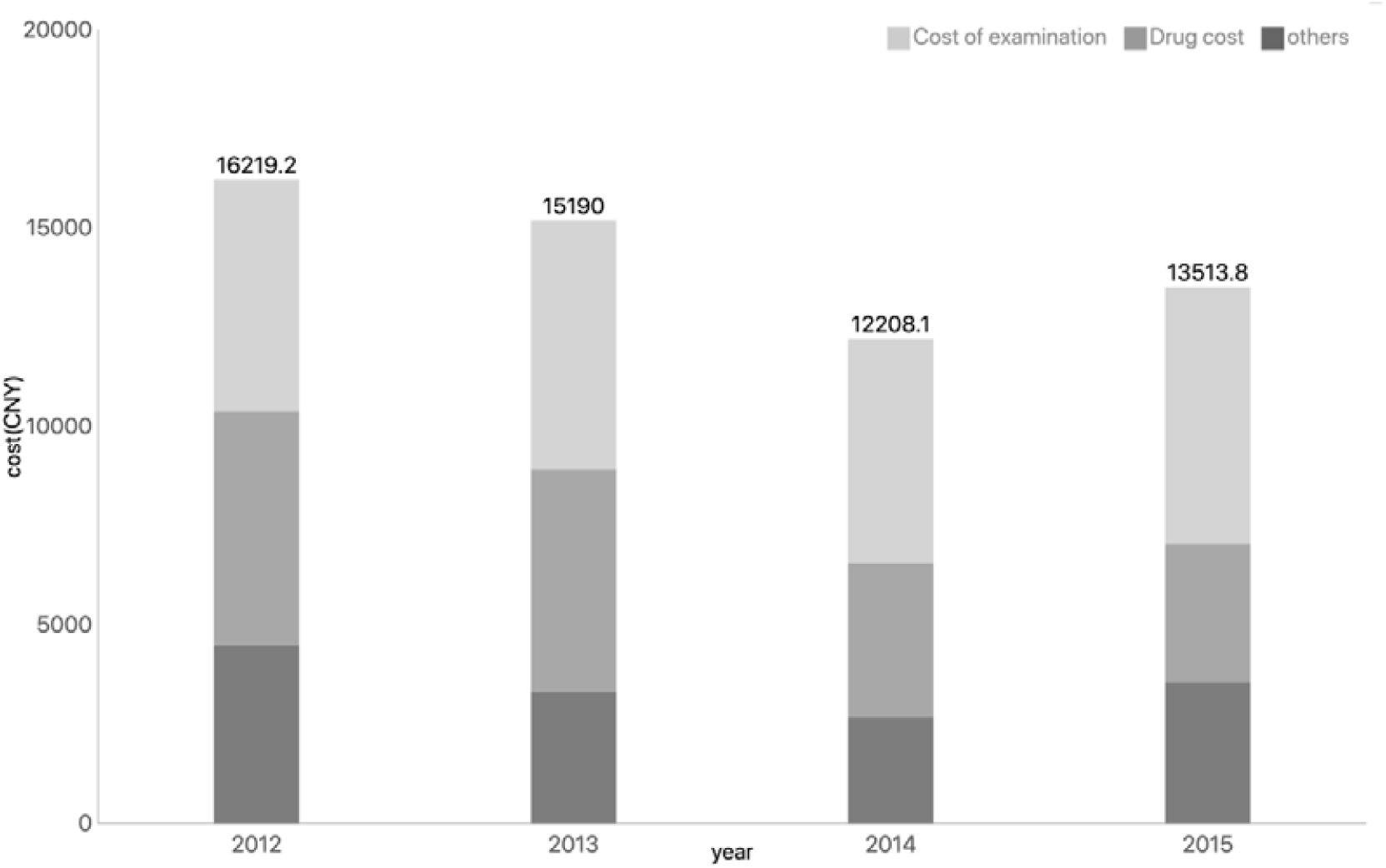
The direct medical cost of hospitalized patients with IPF(2012-2015)

### Regression analysis for the direct medical costs

Univariate regression analysis showed that length of hospital stay, emergency treatment, ventilator use and being a Beijing native were significantly (P<0.05) associated with direct medical costs (Table 6). Multivariate regression analysis showed that the length of hospital stay had the biggest impact. The direct medical costs were not significantly associated with gender or age.

**Table 6.**
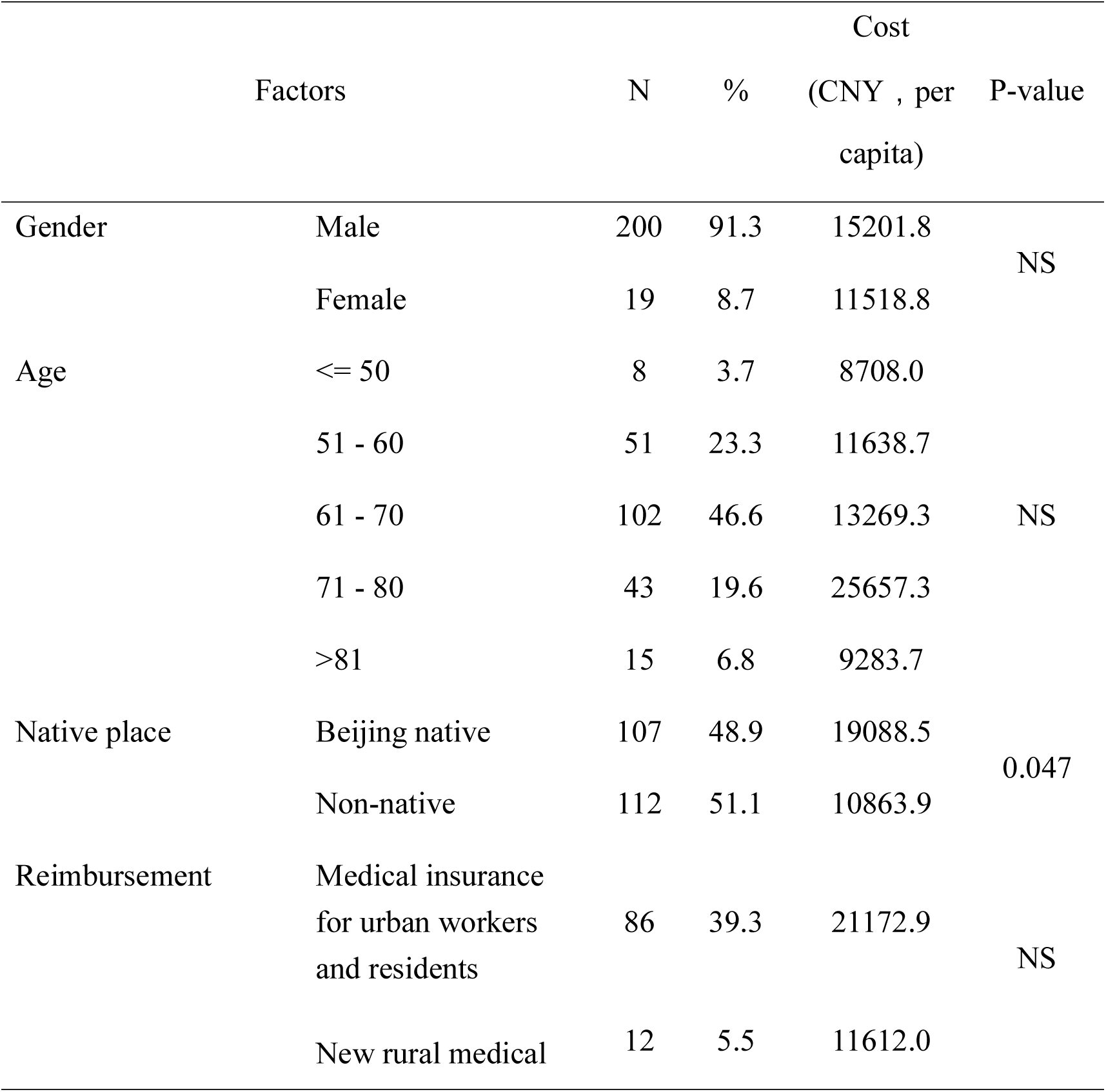
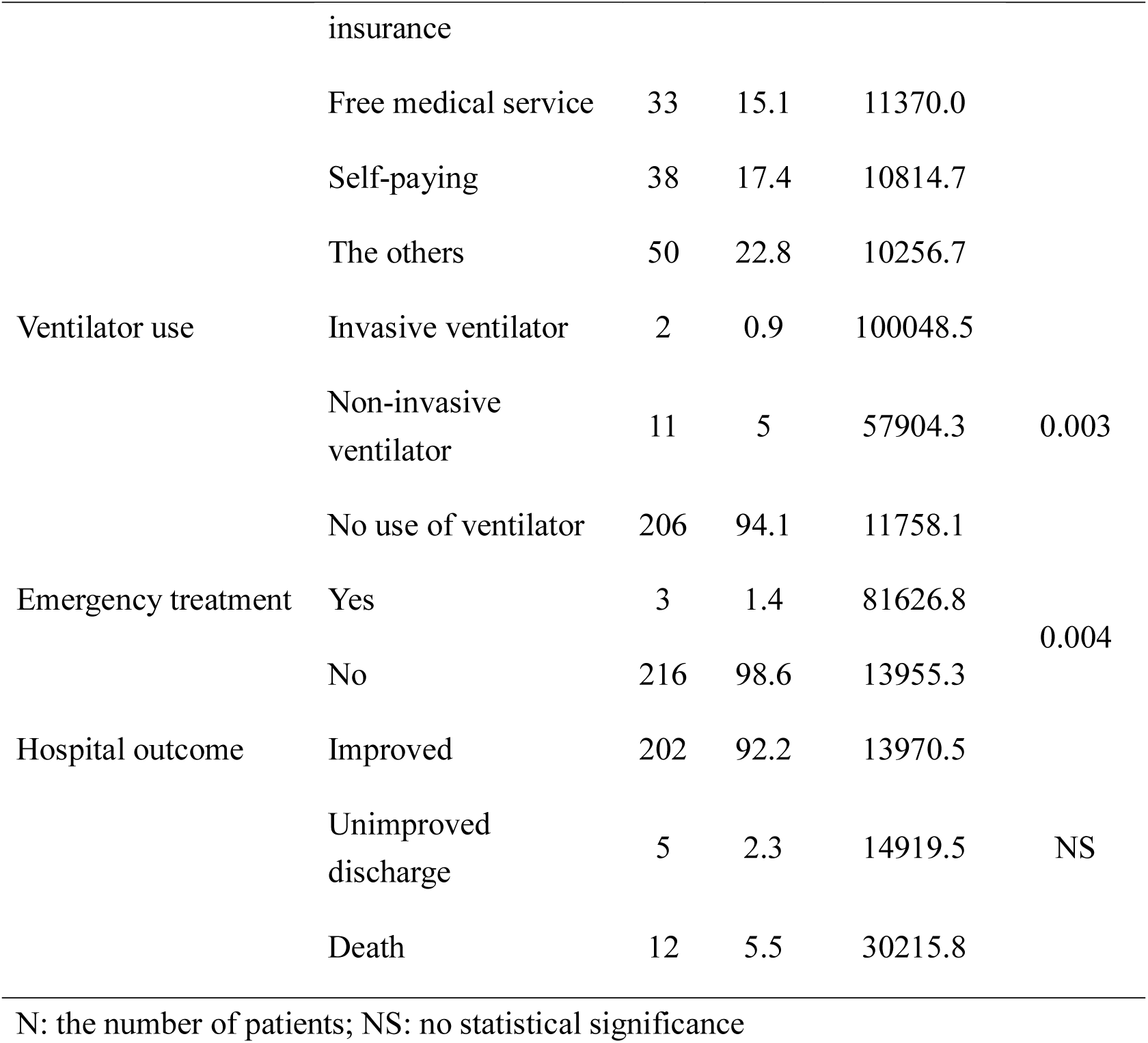
Univariate regression analysis for the direct medical costs

**Table 7.** Multivariate regression analysis for the direct medical costs

| Factors | Unstandardized coefficients |  | Standardized coefficients | T | P-value |
| --- | --- | --- | --- | --- | --- |
|  | B | SE | Beta |  |  |
|  | 8.585 | 0.044 |  | 194.186 | 0.000 |
| Length of stay | 0.060 | 0.004 | 0.707 | 16.890 | 0.000 |
| Emergency treatment | 1.122 | 0.311 | 0.193 | 3.602 | 0.000 |
| Ventilator use | 0.258 | 0.130 | 0.110 | 1.990 | 0.048 |

### Analysis of co-morbid conditions and the direct medical costs

Co-morbid conditions with a significant impact on the direct medical costs included respiratory failure (P=0.004), emphysema (P=0.033), HBP (P=0.026). For the patients with pulmonary arterial hypertension, the cost increased significantly, with P value less than 0.1. The impact on direct medical costs was greatest for respiratory failure (30898.3 CNY), being 3-fold higher than in patients without respiratory failure, followed by the direct medical costs for those with pulmonary arterial hypertension (almost 3-fold increased). (Table 8, Fig 2).

**Table 8.** Analysis of co-morbid diseases and the direct medical costs

| Co-morbid diseases |  | Cost (CNY , per capita) |  | P-value |
| --- | --- | --- | --- | --- |
|  |  | Mean | SD |  |
| Pulmonary infection | Yes | 23480.2 | 48961.6 | 0.121 |
|  | No | 10843.0 | 15630.1 |  |
| Respiratory failure | Yes | 30898.3 | 61084.1 | 0.004 |
|  | No | 10969.3 | 14877.4 |  |
| Pulmonary arterial hypertension | Yes | 26898.2 | 63058.5 | 0.098 |
|  | No | 12278.9 | 16915.5 |  |
| Heart failure | Yes | 16234.8 | 19494.9 | 0.383 |
|  | No | 14824.3 | 31403.7 |  |
| Lung cancer | Yes | 8806.1 | 3900.9 | 0.586 |
|  | No | 14995.3 | 31249.3 |  |
| Emphysema | Yes | 25368.3 | 70895.7 | 0.033 |
|  | No | 12887.7 | 13570.3 |  |
| Asthma | Yes | 8287.1 | 2844.1 | 0.585 |
|  | No | 15036.4 | 31318.2 |  |
| Bronchiectasis | Yes | 14581.6 | 4206.0 | 0.770 |
|  | No | 14886.5 | 31188.5 |  |
| HBP | Yes | 24659.4 | 58634.6 | 0.026 |
|  | No | 11523.3 | 8919.5 |  |
| Diabetes | Yes | 11635.6 | 10979.1 | 0.415 |
|  | No | 15479.0 | 33359.8 |  |
| Coronary artery disease | Yes | 22547.6 | 61584.3 | 0.321 |
|  | No | 13116.7 | 17547.7 |  |

**Fig. 2.**
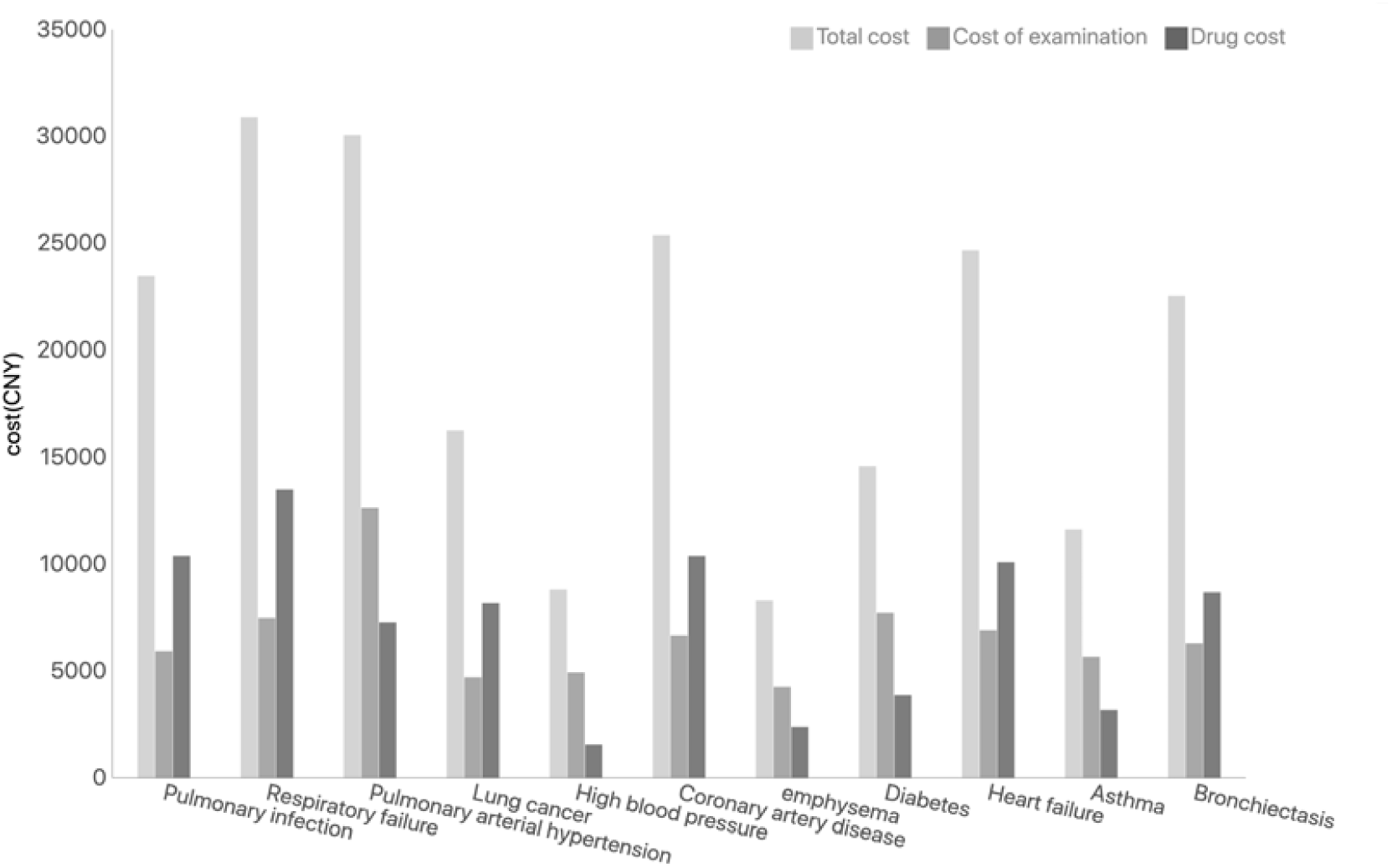
The direct medical cost of IPF in the presence of co-morbid conditions

### Relationship between lung function and costs

131 cases completed lung function tests while in hospital. Analysis of FVC, FVC% pred, DLCO, and DLCO% pred showed that FVC and FVC% pred were negatively correlated with their direct medical costs (P < 0.05). Moreover, the total drug costs increased significantly with increasing severity of diffusion impairment (Table 9).

**Table 9.** Relationship between lung function and costs of IPF patient (CNY, per capita) DLCO% pred: Carbon monoxide diffusion as a percentage of the predicted value *:The costs were significantly different

| DLCO% pred |  | Case ( % ) | Total cost | P-value | Drug cost* | P-value | Cost of examination | P-value |
| --- | --- | --- | --- | --- | --- | --- | --- | --- |
| Normal | ≥80% | 5 ( 4% ) | 7491.8 |  | 756.6 |  | 4883.6 |  |
| Mild | 60-80 % | 21 ( 16% ) | 8839.2 |  | 768.1 |  | 6046.4 |  |
|  |  |  |  | 0.65 |  | 0.03 |  | 0.95 |
| Moderate | 40-60 % | 35 ( 27% ) | 9150.3 |  | 1183.8 |  | 6234.3 |  |
| Severe | < 40% | 70 ( 53% ) | 9784.9 |  | 1916.8 |  | 6059.4 |  |

## Discussion

This study investigated health care resource utilization and the direct medical costs of patients with IPF, from a single center database representing medical claims and health-care costs. We found that there were 219 patients diagnosed with IPF from 2012 to 2015.

The age span of IPF patients was 41-88 years old, with an average age of 65 years old and a male predominance, as previously reported.[8-10]Most importantly, we found that the direct medical economic burden of the IPF inpatients was 14882.3 CNY per patient per admission, of which the costs of examination accounted for 41% of the total, as the largest part, which was similar to a recent study in china.[11]The serious lung function decrease, having comorbities such as respiratory failure, emphysema and high blood pressure, accompanied by length of hospital stay, emergency treatment, ventigator use were the main factors related to increased costs.

COPD is a disease with a huge economic burden. In China, mean annual direct medical costs have been estimated at around 24,372 CNY for a patient with COPD.[12]A Danish study showed that the annual net costs were €8572 for a patients with COPD, which was only one third of that of patients with IPF.[13] In the US, total direct medical costs (including inpatient services, outpatient services and medication claims) and inpatient costs for patients with IPF were found to be $26,378 and $9,100 per person-year, respectively, approximately 2-times higher than controls.[14]In Spain, the estimated annual costs per IPF patient with stable disease, slow and rapid disease progression was €11,484, €20,978 and €57,759, respectively. This corresponded to a weighted average annual cost of €26,435.[15]The costs of our study was calculated from only a single hospitalization of patients with IPF during 2012 to 2015, which was more than half of the annual costs for a Chinese patient with COPD. When the medical costs of whole year including the outpatient services would be calculated together, the total costs for IPF would be much higher. In 2015, the average per capita adjusted net national income in the USA was $ 48,967, while it was only $ 6,352 in China.[16] All these suggested that IPF was also disease condition resulting in large economical burden in China.

Most of patients were first diagnosed as IPF following the approach of ILD during their hospitalization, so the examination fees ranked first, followed by the drug fees, accounting for one third. The Spanish study also found that a significant increase in the annual cost per patient was due to the treatment of acute exacerbation.[15]Nowadays, the treatment options of IPF are limited to the internationally recommended antifibrotic drugs pirfenidone and nintedanib.[17-19]However the costs for these drugs were not relevant to this study, because pirfenidone has not been supplied by the hospital pharmacy at the time of this study, and nintedanib was not available in China until 2018. The estimated total cost had approxiamately 5 times increase to around €80,000 after use of pirfenidone and nintedanib in France.[20]So the economical burden would be increased with the use of antifibrotic drugs.

Regarding the impact of factors related to the direct medical costs, the regression analysis found that length of stay, emergency treatment, ventilator use and Beijing residency were significantly associated with total hospitalization costs. Among these factors, the length of hospital stay had the biggest impact. We know that a variety of factors affect the duration of hospitalization, including disease severity. Thus, the rational use of the allocation of medical resources can significantly reduce the direct medical economic burden on patients. In this study, 5% and 1.4% patients had ventilator use and emergency treatment during the hospitalization, which would consume more health care resources and need more complex medical therapies, inducing more costs.

The patients with IPF often have complications and other comorbidities which include pulmonary arterial hypertension, emphysema, diabetes, lung cancer, GERD, and cardiovascular disease [3,21] and require substantial health care resources, leading to increased overall burden of illness.[22]Ning Wu found that patients with IPF have a high burden of co-morbid conditions and HRU compared to non-IPF patients.[23]Collard et al found that pulmonary infection, coronary artery disease, diabetes and heart failure were the most prevalent comorbidity and all were significantly more common in IPF than in controls.[14]In this study, which was a cross-sectional review, there were only 4 patients with lung cancer in IPF and only 19 patients completed the 24-hour ambulatory esophageal pH and pressure recording. As a result, the GERD and lung cancer prevalence was lower than the previews study and the group was too small to be statistically significant.[24,25]As our study showed, pulmonary infection, high blood pressure, coronary artery disease, respiratory failure, diabetes, emphysema and pulmonary arterial hypertension were the most prevalent comorbidity codes in the IPF. Another study in china had showed that the prevalence of IPF patients with pulmonary arterial hypertension and emphsema was 29% and 42% respectively.[26]The costs of IPF patients with respiratory failure and pulmonary arterial hypertension were found to be higher than for other patients. This results are similar to a previous study.[27]Most series have shown a higher mortality when pulmonary arterial hypertension is present in IPF patients.[28-29]Thus, an IPF patient with pulmonary arterial hypertension will raise the economic burden.

There are several limitation. firstly, this is a retrospective research from a single center. Secondly, only the costs for hospitalization and not annual costs were analyzed. Thirdly, the costs of antifibrotic drugs were not included due to lack of available drugs during the study time.

## Conclusion

As the prevalence of IPF appears to be rising along with an increasing burden on our healthcare system, a substantial increase in public awareness and research funding will be necessary to address the unmet needs and reduce the clinical and economic burden of this still incurable illness.

## Data Availability

Data sharing statement: The data sets used for this study are available

## Acknowledgments

We would like to acknowledge the medical record room and statistics office in Beijing Chaoyang Hospital for providing us with data support. Thanks also to our entire team for their assistance in data collection and processing. At the same time, I would like to thank the DR. Costabel for guiding the article. At last but not least, the authors would like to thank study participants for their time, patience and involvement in the study.

## Notes

Funding: Supported by National Key Technologies R & D Program Precision Medicine Research (No.2016YFC0901101), CAMS Innovation Fund for Medical Sciences (CIFMS, No. 2018-12M-1-001) and Non-profit Central Research Institute Fund of Chinese Academy of Medical Sciences (No. 2019PT320021)

### Competing Interest Statement

The authors have declared no competing interest.

### Funding Statement

Supported by National Key Technologies R & D Program Precision Medicine Research (No.2016YFC0901101), CAMS Innovation Fund for Medical Sciences (CIFMS,No. 2018-12M-1-001)and Non-profit Central Research Institute Fund of Chinese Academy of Medical Sciences (No. 2019PT320021)

### Author Declarations

All relevant ethical guidelines have been followed and any necessary IRB and/or ethics committee approvals have been obtained.

Any clinical trials involved have been registered with an ICMJE-approved registry such as ClinicalTrials.gov and the trial ID is included in the manuscript.

